# Evaluating the clinical performance of a novel dual-target stool DNA test for colorectal cancer detection

**DOI:** 10.1101/2021.03.10.21252951

**Authors:** Zhongxin Wang, Jian shang, Guannan Zhang, Lingjun Kong, Feng Zhang, Ye Guo, Yaling Dou, Jun Lin

## Abstract

**BACKGROUND AND AIMS:** Methylation-based stool DNA test showed a promising application for colorectal cancer (CRC) detection. This study aimed to evaluate the performance of a novel dual-target stool DNA test (DT-sDNA), composed of *SDC2* and *TFPI2*, for the detection of CRC in clinical practice by using large-scale data from a multicenter clinical trial.

**METHODS:** We enrolled 1,164 participants from three independent hospitals, including 320 CRC patients, 148 adenomas, 396 interfering diseases and 300 healthy controls. Their stool samples were collected and tested paralleled by DT-sDNA test under the guidance of standard operating procedure. All participants were dichotomized as positive and negative according to the cycling threshold (Ct) values measured by quantitative methylation-specific PCR (MSP). The diagnostic performance of DT-sDNA test was assessed by calculating indexes of sensitivity, specificity, and overall coincidence rate. Sanger sequencing and retesting of resected participants were performed to verify the accuracy and effectiveness of the test.

**RESULTS:** Overall, the sensitivity, specificity, and total coincidence rate of DT-sDNA test for CRC detection were 95.31%, 88.39%, and 90.29%, respectively, with the kappa value of 0.775 (P < 0.05) when comparing to non-CRCs. The sensitivities for the detection of advanced adenomas (n=38) and non-advanced adenomas (n=110) were 63.16% and 33.64%, and the specificity was 96.67% for healthy normal controls. The methylation status of *SDC2* and *TFPI2* in 375 samples were verified by Sanger sequencing and the average coincidence rate reached 99.62%. The coincidence rate was 94.12% (n=32) for 34 participants that undertook DT-sDNA test again after surgical resection. These results demonstrated high accuracy of the DT-sDNA test in discriminating CRCs from other diseases and healthy controls.

**CONCLUSIONS:** The novel DT-sDNA test showed good performance for the diagnosis of colorectal cancer in clinical practice; fda.hubei.gov.cn number; 20190020787.

## INTRODUCTION

Colorectal cancer (CRC) is one of the most common digestive cancers. Both the incidence and mortality rates of CRC ranked as the top five among all cancers in China ^1^. A lower CRC incidence was found in China than that in USA and UK, whereas the mortality was much higher ^2^, which might be related to the lower rate of early detection in Chinese. Several screening methods were developed for the diagnosis of CRC and each of them has individual features that can affect the perceptions and preferences of both patients and physicians. Colonoscopy is a highly specific test for CRC and thus is recommended as the reference method for CRC screening with high sensitivity. However it has disadvantages of invasiveness and low patient compliance ^3^. Computed tomography (CT) colonography shows high sensitivity and specificity in detecting both tumors and polyps, but has limitations such as invasive and radiation exposing ^4^. Guaiac fecal occult blood test (gFOBT) and fecal immunological test (FIT) are two methods based on stool samples with obvious advantages of noninvasiveness and convenience ^5^, while they showed lower sensitivities due to the reason that not all patients with CRC had fecal occult blood ^6^.

Multi-target stool DNA (MT-sDNA) test based on abnormally methylated DNA is an option for the screening of asymptomatic persons at average risk of CRC. Several MT-sDNA tests have been recently evaluated for their use in CRC detection and the associated results demonstrated their superior sensitivity during clinical practices, which is almost closed to the reference method of colonoscopy ^7–9^. Cologuard® (Exact Sciences Corporation, USA) was the first commercially MT-sDNA test approved by US Food and Drug Administration (FDA) for using population-wide to screen average-risk individuals. This MT-sDNA test detected two methylated DNA biomarkers and the mutations of KRAS, as well as hemoglobin biomarker simultaneously. Its clinical sensitivity achieved 92.3% which is superior as compared to FIT although the specificity was a little inferior ^7^. Unlike Cologuard®, the commercial products of “EarlyTect-Colon Cancer” (Genomictree, South Korean) and Colosafe (Creative Biosciences CO., Ltd., China), approved by Korean Food and Drug Administration and China National Medical Products Administration respectively, only detect methylated SDC2 as target ^9, 10^. Single-target stool DNA (ST-sDNA) tests have the advantages over MT-sDNA test such as only fecal sample is required and are more patient compliance. However, the reported sensitivities for ST-sDNA tests are much lower than MT-sDNA tests which almost does not exceed 90% ^11^. The approved Epi proColon 2.0 assay by USFDA and Chinese FDA (CFDA) is a blood-based test that detects aberrant methylated *SEPT9* in CRCs ^12^. Its ability to detect CRC varied greatly in different studies with a pooled sensitivity of 72% and specificity of 92% ^13^. Furthermore, the sensitivity for adenomas and polyps was only 0.15 making it unsuitable for screening test.

Several types of research have reported frequently hypermethylated status of SDC2 and TFPI2 in tumor tissues of CRC, but rarely in normal colon tissues ^14–19^ and that was confirmed in our preliminary studies (data prepared for publishing). In this multicenter, double-blinded case-control study, the clinical performance of the DT-sDNA test was comprehensively evaluated in more than 1,000 participants. In addition, the association of the test with patient’s clinical characteristics was also analyzed. These outcomes will provide robust support for the early detection of colorectal cancer in clinical practice.

## MATERIALS AND METHODS

### Study Design

In this study, more than 1,000 individuals were enrolled from three hospitals (Peking Union Medical College Hospital, The First Affiliated Hospital of Anhui Medical University, and Zhongnan Hospital of Wuhan University) under the Technical Guidelines for Clinical Trials of in vitro Diagnostic Reagents (CFDA notice No. 16 2014). According to the guidance of In-Vitro Diagnostic Reagent Registration released by China Food and Drug Administration (2014), this clinical trial was registered in Hubei Food and Drug Administration, China (NO. 20190020787, http://fda.hubei.gov.cn/bsfw/bmcxfw/bjcx/). The study was performed parallelly in three centers after it was approved by the institutional ethics review committee of each hospital. The approved ID numbers of Peking Union Medical College Hospital, The First Affiliated Hospital of Anhui Medical University, and Zhongnan Hospital of Wuhan University were KS2020260, PJ2019-09-04, and 2019009-1K, respectively. All participants signed informed consent and were told the test results. The primary measures of this study, including sensitivity, specificity and total coincidence rate, were calculated to evaluate the accuracy of this DT-sDNA test as compared with the reference method of colonoscopy or pathology.

### Participant enrollment

This study enrolled 1,196 participants in three hospitals from July 2019 to May 2020. All participants were over 18 years old. The included CRC patients must meet the following qualifications: 1) the disease status was confirmed by colonoscopy or histopathological examination, and 2) did not undertake resections and 3) no radiotherapy or chemotherapy. Interfering diseases include other untreated digestive tumors (stomach, liver, esophagus, and pancreatic cancer) and benign digestive diseases (esophagitis, gastritis, enteritis, inflammatory bowel disease, appendicitis, peptic ulcer, colorectal polyps, colorectal adenoma, and hemorrhoids). The recruited participants were then excluded if their stool DNA samples or the Ct values measured by MSP did not pass quality controls.

### Samples Preparation

About 10∼15g fresh fecal samples from all participants were collected before surgical resection and then transferred to a 45 ml storage tube that contained 30 ml of preservative buffer (Wuhan Ammunition Life-tech Company, Ltd., Hubei, China) within four hours. All the collected fecal samples were assigned a unique id for blinded testing before they were delivered to the medical testing laboratory of each center or stored at −80 °C until use if they were not processed in time. The corresponding clinical characteristics of patient age, gender, TNM stage ^20^, anatomical location, and differentiation were also collected. Patients with the anatomical location of cecum, ascending colon, transverse colon, and hepatic flexure were grouped as right-sided tumors, while for descending colon, sigmoid colon, and rectosigmoid junction, they were classified as left-sided tumors. Two serum biomarkers, CEA and CA19-9 were retrieved for following analysis if they were available.

### Bisulfite Treatment and Methylation Specific PCR

Stool DNA was isolated from fecal samples using the stool DNA extraction kit (Wuhan Ammunition Life-tech Company, Ltd., Hubei, China) according to the instructions. The purified DNA was then treated with sodium bisulfite to convert unmethylated cytosine to uracil while leaving methylated cytosine unchanged. Briefly, the purified DNA, conversion buffer and protection buffer were incubated at 98°C for 10 min, and then treated with sodium bisulfite at 64°C for 1h. The bisulfite-treated DNA was then centrifuged at 12000rpm for 30 seconds for purifying. After two steps of washing, the DNA was dried at room temperature and eluted with 40ul TE buffer.

The converted stool DNA was tested by DT-sDNA test kit developed by Wuhan Ammunition Life-tech Company, Ltd under the instructions. The methylation status of *SDC2* and *TFPI2* were measured by Ct values of MSP. *ACTB* was used as an internal MSP control and a measurement was considered valid only if the Ct value of *ACTB* was no more than 34. An invalid measurement would be performed second DNA extraction, bisulfite conversion, and MSP. Samples were assigned negative detection if both of the Ct values of *SDC2* and *TFPI2* were larger than 38, otherwise, they were assigned positive detection. All the undetermined Ct values of *SDC2* and *TFPI2* were arbitrarily given a value of 45 for the largest Ct is 45 in ABI 7500 real-time PCR device.

### Outcome Measures and Statistical Analysis

The primary endpoints were the sensitivity, specificity, and overall coincidence rate. These indexes were calculated by following equations:

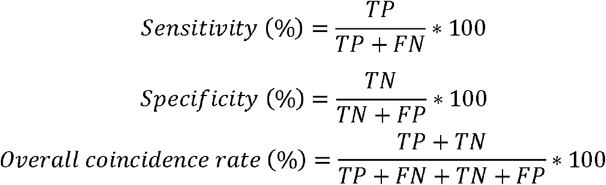

where TP, FN, TN and FP represent true positive, false negative, true negative and false positive. Cohen’s kappa test was used to measure the diagnostic reliability of DT-sDNA test as compared with the reference methods (colonoscopy or histopathological examination). Pearson’s chi-squared test was conducted to determine the difference between two categories. P values < 0.05 were considered as statistically significant. All statistical analysis and figure plotting were performed using R software (v3.6.1). The method and result of statistical analysis were revised by Professor Changming Lu, the principal academic expert of Wuhan Ammunition Life-tech Company, Ltd.

## RESULTS

### Subject Features

After 27 participants that did not meet the inclusion criteria and the other 5 whose DT-sDNA test did not pass the quality control were excluded, we finally included 1,164 participants (**Figure 1**). Of the 1,164 participants, 455 (39.09%) undertook the examination of colonoscopy, while the rest 710 (60.91%) participants received histopathological examinations (**Supplementary Table 1**). The study population consisted of 300 healthy normal controls, 396 participants with interfering diseases, 148 adenomas, and 320 CRCs with the median ages ranging from 49 to 63 years old (**Table 1**). No significant difference in gender distribution was observed in the four aforementioned categories (χ^2^ = 2.26, P = 0.52). CRCs were classified I to IV according to the TNM staging system ^20^, of which 89.69% were stage I-III (**Table 1**). Sanger sequencing were performed for 375 samples that contained 263 and 112 samples whose detected results were consistent and not consistent with their diagnosed disease status to verify the accuracy of DT-sDNA test. Besides, 34 patients were followed up and their stool samples were retested with DT-sDNA test after surgical resections.

**Table 1.** Subject features.

| Clinical feature | Normal | Adenoma | Interfering diseases | CRC |
| --- | --- | --- | --- | --- |
| **Age** |  |  |  |  |
| (median[interquile]) | 49.00(39.00~58.25) | 58.00(50.75~64.25) | 49.00(37.00~57.25) | 63.00(54.00~71.00) |
| **Gender (%)** |  |  |  |  |
| Male | 154 (51.33) | 83 (60.14) | 216 (54.55) | 184 (57.50) |
| Female | 146 (48.67) | 65 (47.10) | 180 (45.45) | 136 (42.50) |
| **Stage (%)** |  |  |  |  |
| I |  |  |  | 49 (15.32) |
| II |  |  |  | 114 (35.62) |
| III |  |  |  | 124 (38.75) |
| IV |  |  |  | 20 (6.25) |
| Not available |  |  |  | 13 (4.06) |
| **Summary (%)** | 300 (25.77) | 148 (12.72) | 396 (34.02) | 320 (27.49) |

**Figure 1.**
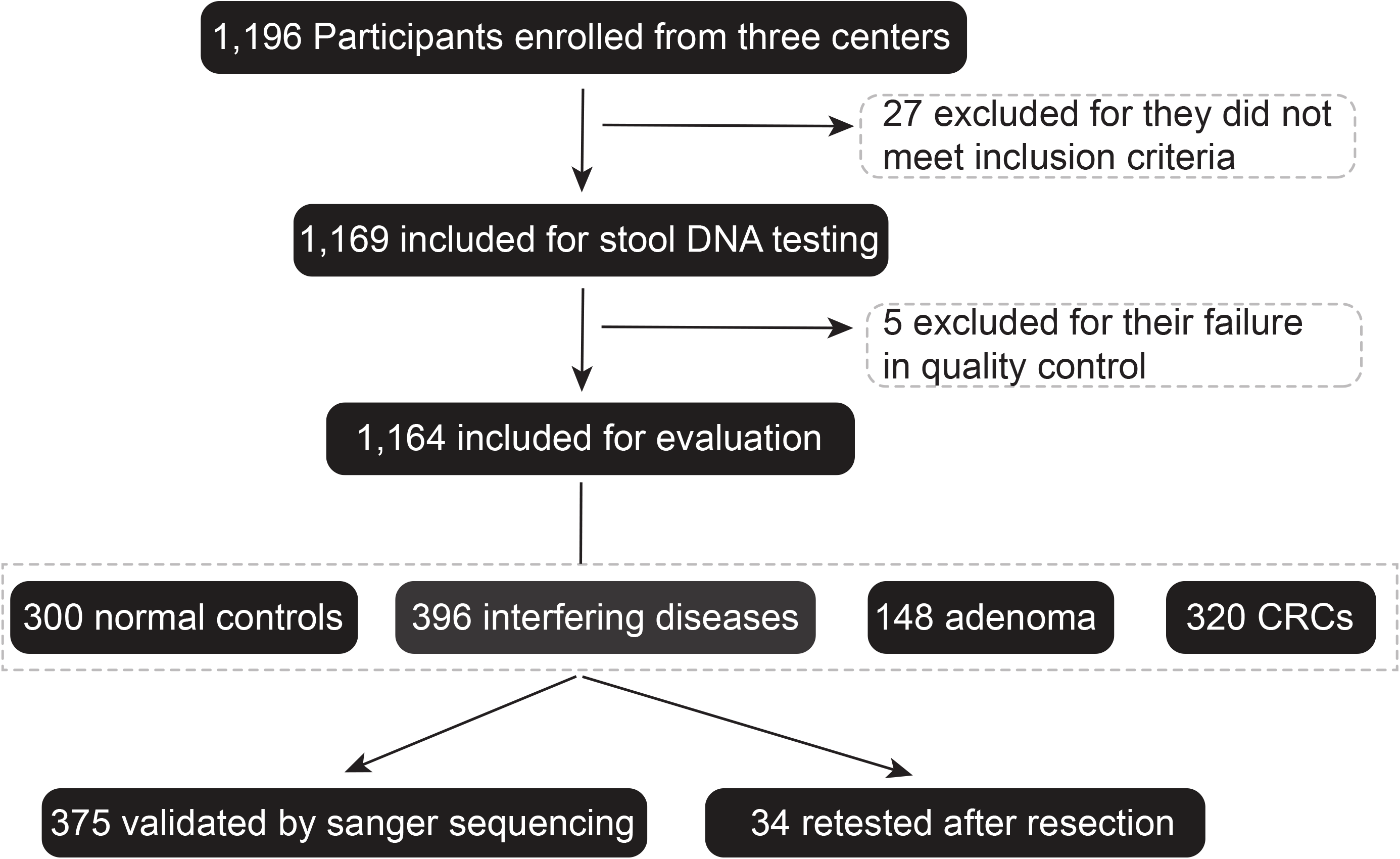
Overview of the clinical trial design. CRC indicates colorectal cancer. The 148 adenoma patients includes 38 advanced adenoma patients and 110 non-advanced adenoma patients.

### Methylation Status of SDC2 and TFPI2 in Study Population

Three independent batches of tests were performed by using 10 positive references (P1-P10) and 10 negative references (N1-N10) to assess the stability of this DT-sDNA test. The amplification curves of MSP showed apparently distinct Ct values for methylated and un-methylated targets (**Figure 2A-D**), indicating that MSP can be used to discriminate positive and negative samples. Good consistency of Ct values of *ACTB* was observed among three independent batches for both positive and negative references (**Figure 3A**) with all Ct values no more than 34 which reflected a robust quality control of the test. Similar results were also obtained for the 100% consistency among the measured results of three independent batches for all 20 references (**Figure 3B**). These results suggested the high stability of the DT–sDNA test.

**Figure 2.**
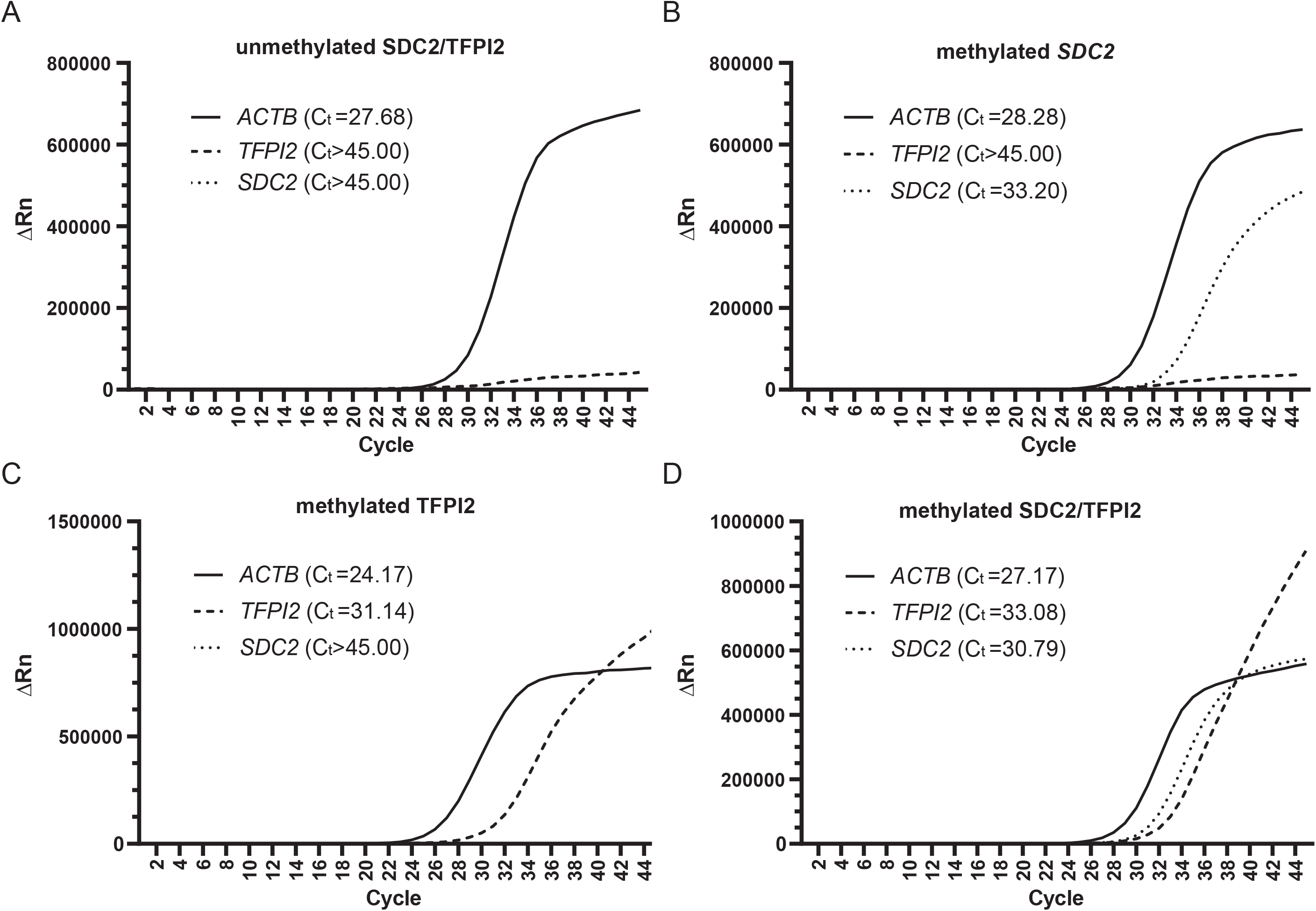
Amplification curves of methylation specific PCR for unmethylated *SDC2/TFPI2* (A), methylated SDC2 (B), methylated TFPI2 (C) and methylated *SDC2/TFPI2*. **(D)**. ΔRn = Rn (post-PCR read) – Rn (pre-PCR read), where Rn = normalized reporter. The X-axis indicate PCR cycles from 1 to 45. Ct values were calculated according to the PCR cycle number at which the fluorescence meets the threshold in the amplification plot.

**Figure 3.**
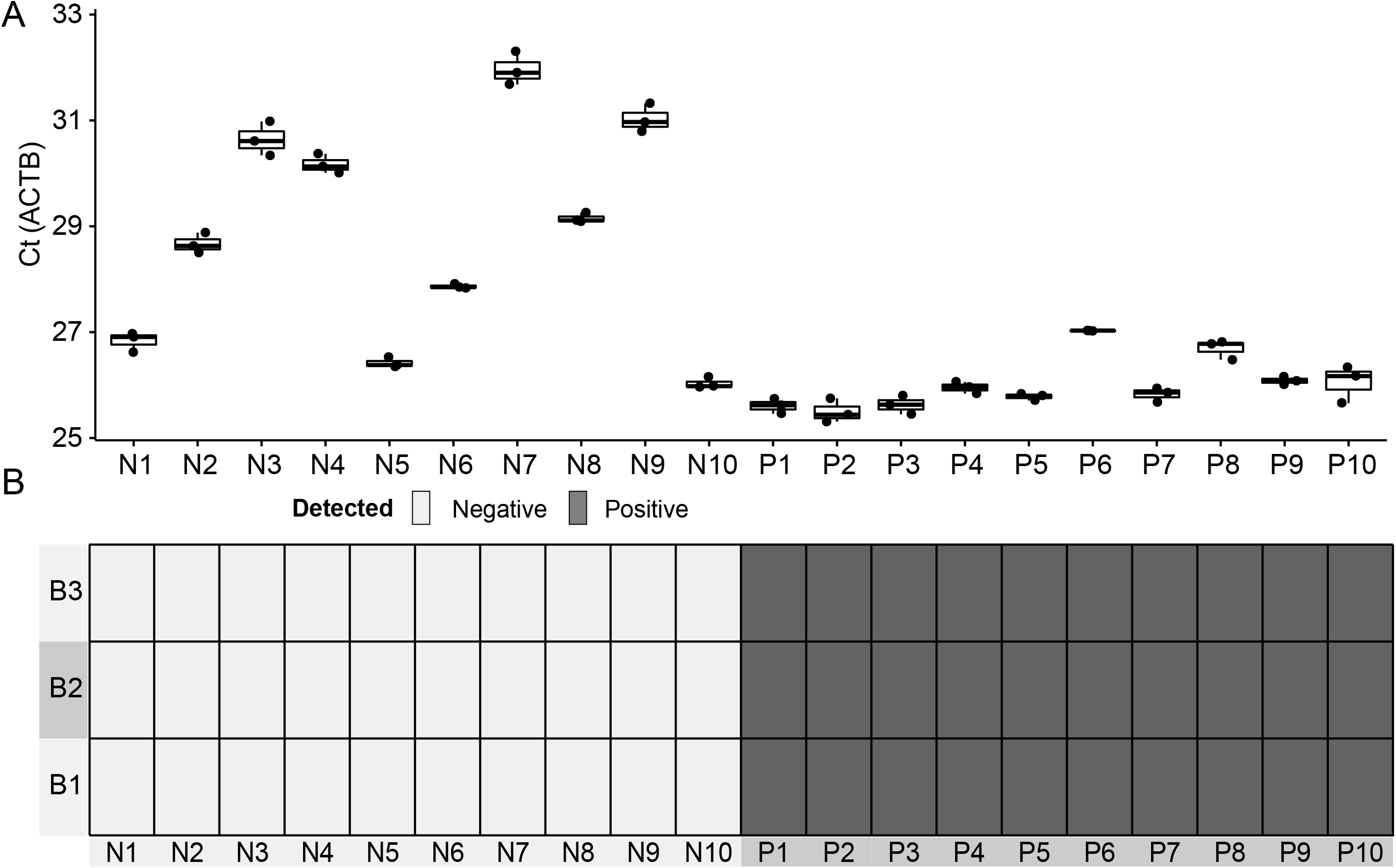
Three batches of DT-sDNA tests for 10 positive and 10 negative references. **A**: Ct values of *ACTB* measured by MSP for 20 references. **B:** Detected results of the 20 references in three batches. N1-N10: 10 negative references, P1-P10: 10 positive references. B1-B3 represents batch 1, 2 and 3.

The methylation profiles of *SDC2* and *TFPI2* were then evaluated among the 1,164 participants by DT-sDNA test. Methylated *SDC2* and *TFPI2* were detected in 261 (22.42%) and 362 (31.10%) samples respectively, and 403 (34.62%) samples contained either methylated *SDC2* or methylated *TFPI2*. We found that the combined sDNA test detected more positive samples, however, 88 (7.56%) samples which were classified as interfering diseases (n=27) and adenomas (n=61) were tested positive (**Figure 4**). Besides, 15 CRCs showed unmethylated *SDC2* and *TFPI2*, which might imply a specific methylator phenotype of CRCs represented by the methylation status of these two genes.

**Figure 4.**
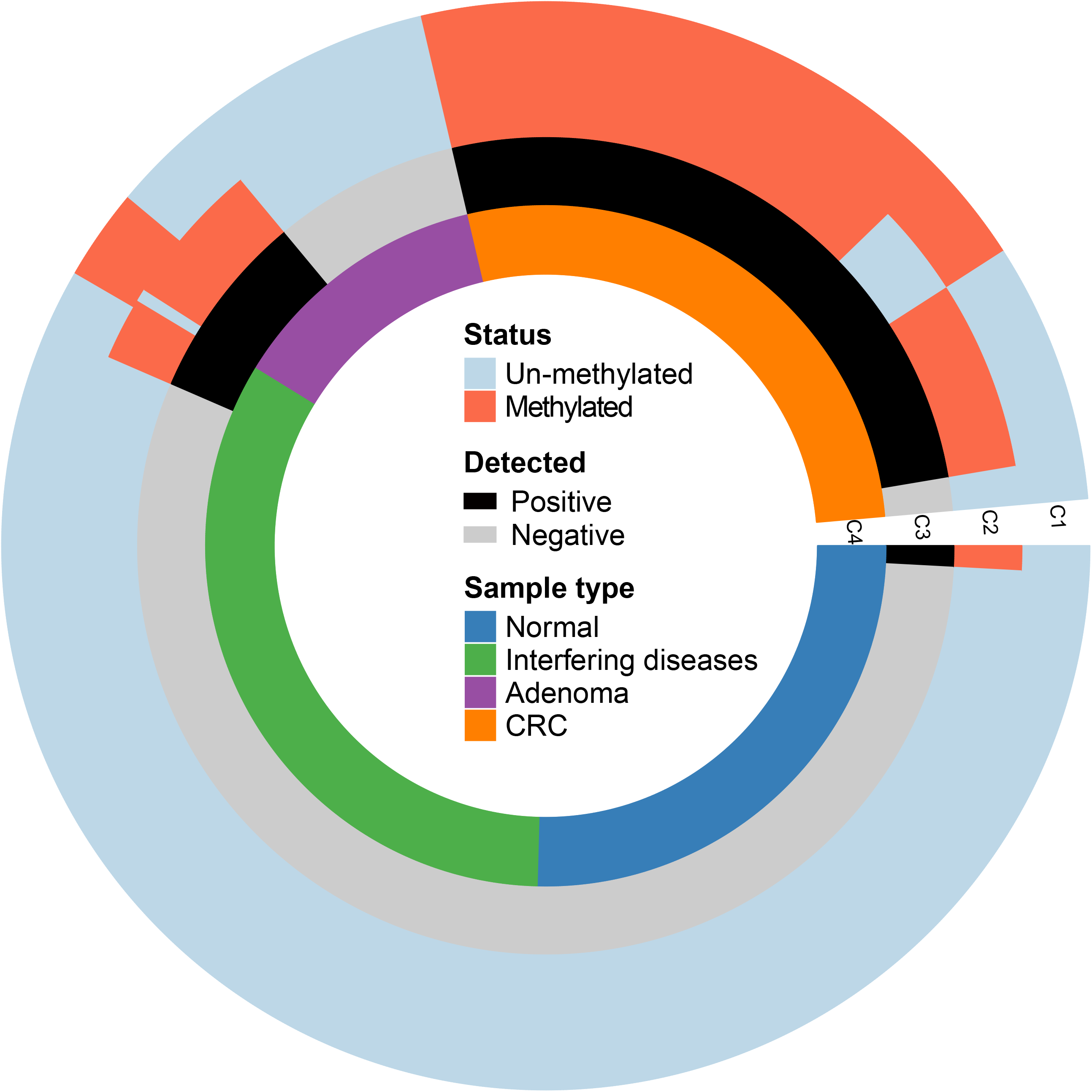
Methylation profiles of *SDC2* and *TFPI2* in study population. C1, C2 indicate the methylation status of *SDC2* and *TFPI2* respectively. C3 represents the detected results of 1,164 enrolled participants. C4 represents four groups of the study population.

### Performance of methylated SDC2 and TFPI2 in Detection of CRCs and Adenomas

The sensitivity, specificity, and area under ROC curve (AUC) were calculated to evaluate the diagnostic performance of DT–sDNA test for CRCs. Overall, the sensitivity for CRC detection was 95.31% (95%CI: 88.64% - 97.21%) with specificities of 88.39% (95%CI: 73.80% - 90.32%) for all non-CRCs and 96.67% (95%CI: 80.49% - 98.35%) for healthy normal controls (**Figure 5A&B**). The AUC was 0.92 (95%CI: 0.90∼0.93) when using all non-CRCs as negative controls, but improved to 0.96 (95%CI: 0.94∼0.97) when compared to healthy normal controls (**Figure 5A&B**). Colorectal adenoma was known as the main precancerous lesion of CRC. We observed a sensitivity of 41.22% (95%CI: 36.36% - 48.68%) for adenoma detection (**Figure 5C**). However, for advanced and non-advanced adenomas, the sensitivities were 63.16% (95%CI: 55.81% - 76.56%) and 33.64% (95%CI: 30.41% - 42.02%) respectively (**Supplemental Table 2**). The sensitivity for early-stage (I and II) CRCs detection was a little higher than that of late-stage (III and IV) CRCs (96.93% vs 93.06%) (**Figure 5D&E**). Additionally, we found that the methylated *SDC2* in detecting both CRCs and adenomas presented higher specificity and lower sensitivity, while it was inversed for the methylated *TFPI2*. This result might elucidate the improved detected performance of the combined DT-sDNA test than these of any target alone.

**Table 2.** Kappa test for agreement of SDC2 and TFPI2 in detection of interfering diseases.

|  |  | Detected Negative | Detected Positive | Kappa (95%CI) | Pr value |
| --- | --- | --- | --- | --- | --- |
| SDC2 | Interfering diseases | 391 | 5 | 0.72 (0.67~0.77) | < 0.001 |
|  | CRCs | 91 | 229 |  |  |
| TFPI2 | Interfering diseases | 371 | 25 | 0.78 (0.73~0.83) | < 0.001 |
|  | CRCs | 51 | 269 |  |  |
| SDC2/TFPI2 | Interfering diseases | 369 | 27 | 0.88 (0.85~0.92) | < 0.001 |
|  | CRCs | 15 | 305 |  |  |

**Figure 5.**
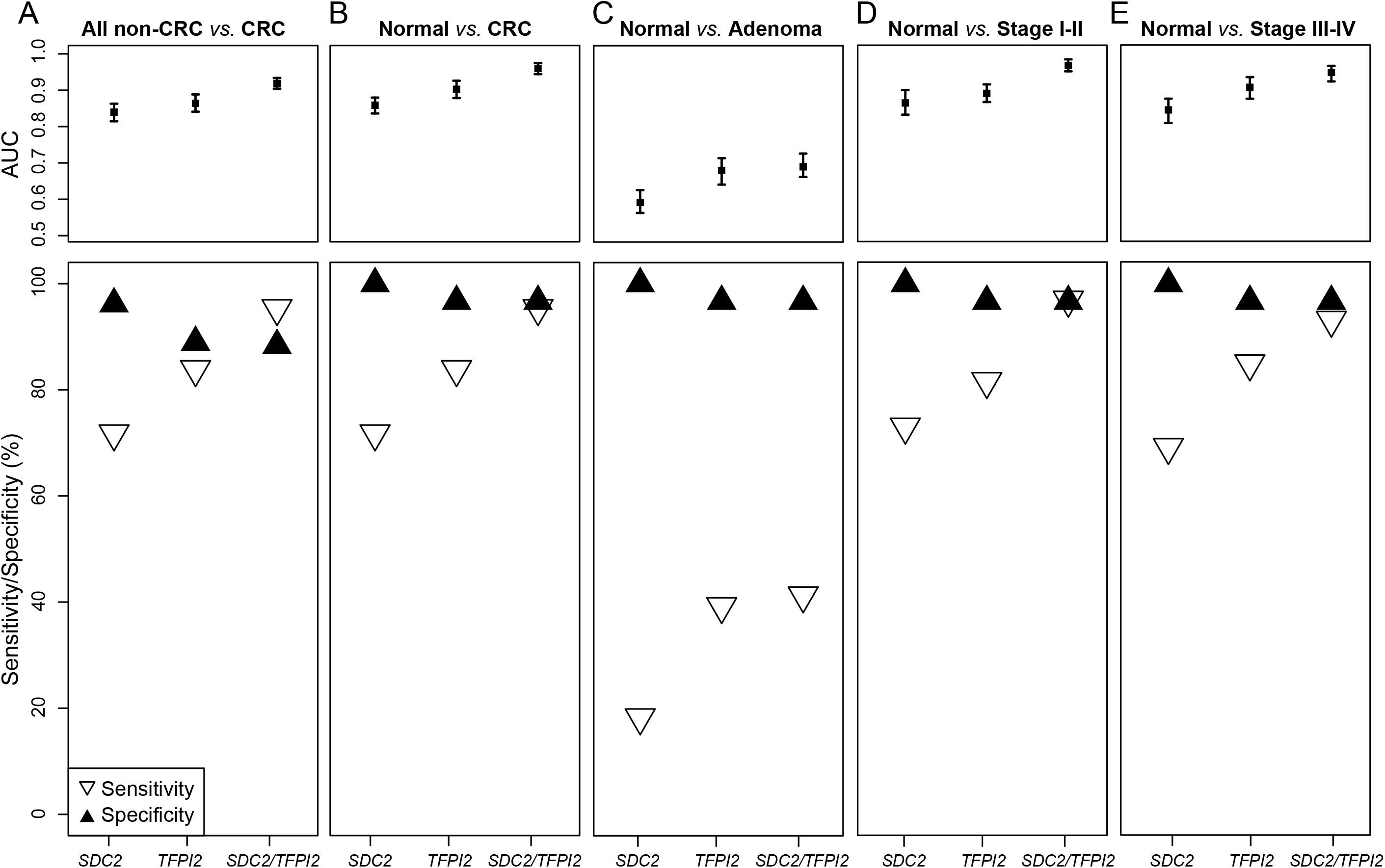
Performance of the SDC2 and TFPI2 methylation in detection of adenoma and CRCs. All non-CRCs include healthy normal controls, interfering diseases, and adenomas. Advanced adenomas and non-advanced adenomas are integrated together as adenomas.

### Performance of methylated SDC2 and TFPI2 in Detection of Interfering Diseases

The study recruited 396 participants who suffered the common digestive diseases and they were classified as interfering diseases. The positive detection rate for interfering diseases was 6.70% (n=27), which referred to the specificity of 93.15% (n=367) (**Supplemental Table 3**). Kappa values for methylated SDC2, TFPI2 and the combined dual-target were 0.72, 0.78 and 0.88 respectively (**Table 2**), suggested a good agreement of DT-SDNA test in discriminating interfering diseases from CRCs.

**Table 3.** Validation by Sanger sequencing.

|  |  | Sanger sequencing |  | Kappa (95%CI) | Pr value |
| --- | --- | --- | --- | --- | --- |
|  |  | Negative | Positive |  |  |
| SDC2 | Negative | 258 | 0 | 1.00 (1.00~1.00) | < 0.001 |
|  | Positive | 0 | 117 |  |  |
| TFPI2 | Negative | 187 | 1 | 0.98 (0.96~1.00) | < 0.001 |
|  | Positive | 2 | 185 |  |  |

### Validation by Sanger Sequencing and Surgical Following

The methylation status of *SDC2* and *TFPI2* in 375 samples were verified by performing Sanger sequencing. The unmethylated cytosine in a CpG site will be transformed to thymine after the treatment of sodium bisulphite, while the methylated cytosine is not changed. By sequencing the products of MSP, we can directly determine whether the target is methylated according to sequence of CpG sites. We observed a 100% consistency of the methylation status of *SDC2* between the DT-sDNA tests and sequenced results (kappa = 1.00) (**Table 3**). The kappa value for *TFPI2* achieved 0.98, despite that 3 samples were not in line between the two methods. Furthermore, 34 CRC patients were followed-up after resected and their stool samples were retested by DT-sDNA test under the same operations. Four CRCs that were recognized as false negative before surgical resection were detected negative again after resection, and the rest 28 CRCs that were positive detections showed negative tests after resection. The overall consistency rate was 94.12% (32/34) except for two CRCs that were disagreed with expected (**Supplemental Table 4**).

**Table 4.** Positive detection rate of DT-sDNA test for different clinical features of CRCs.

| Characteristics | CRCs | Stool DNA test |  |  | X<sup>2</sup> test P |
| --- | --- | --- | --- | --- | --- |
|  |  | SDC2 (% ,n) | TFPI2 (% ,n) | SDC2/TFPI2 (% ,n) |  |
| **Gender** |  |  |  |  | 0.97 |
| Male | 184 | 70.65 (130) | 85.33 (157) | 95.65 (176) |  |
| Female | 136 | 72.79 (99) | 82.35 (112) | 94.85 (129) |  |
| **Age** |  |  |  |  | 0.92 |
| <50 | 54 | 74.07 (40) | 85.19 (46) | 94.44 (51) |  |
| 50~60 | 70 | 70.00 (49) | 81.43 (57) | 94.29 (66) |  |
| 60~70 | 106 | 70.75 (75) | 83.96 (89) | 96.23 (102) |  |
| >=70 | 90 | 72.22 (65) | 84.62 (77) | 95.56 (86) |  |
| **TNM Stage** |  |  |  |  | 0.21 |
| I-II | 163 | 73.01 (119) | 81.60 (133) | 96.93 (158) |  |
| III-IV | 144 | 68.75 (99) | 85.42 (124) | 93.06 (134) |  |
| Not available | 13 | 84.62 (11) | 100 (13) | 100 (13) |  |
| **Location** |  |  |  |  | 0.44 |
| Left-sided | 53 | 69.81 (37) | 90.57 (48) | 96.23 (51) |  |
| Right-sided | 32 | 87.50 (28) | 62.50 (20) | 96.88 (31) |  |
| Other colon | 67 | 62.69 (42) | 76.12 (51) | 92.54 (62) |  |
| Rectum | 151 | 74.17 (112) | 91.39 (138) | 96.69 (146) |  |
| Ileocecal valve | 17 | 58.82 (10) | 70.59 (12) | 88.24 (15) |  |
| **Differentiation** |  |  |  |  | 0.12 |
| High | 24 | 70.83 (17) | 75.00 (18) | 87.50 (21) |  |
| Moderate | 280 | 71.07 (199) | 85.00 (238) | 95.71 (268) |  |
| Low | 16 | 75.00 (12) | 87.50 (14) | 100.00 (16) |  |

### Association of Clinical Features with Methylation Status of SDC2 and TFPI2

The association of DT-sDNA test with patient clinical features was assessed by calculating the detection rate of different groups of CRCs. The positive detection rate of DT-sDNA did not show significant variation in patient’s gender, age, TNM stage, tumor location, and differentiation (**Table 4**). Notably, the positive detection rates of *TFPI2* alone in all categories were relatively higher than these of *SDC2* alone, except for the right-sided, which might be related to the significantly different histological and molecular characteristics between left-sided and right-sided CRCs ^21, 22^. Two serum tumor markers, carcinoembryonic antigen (CEA) and carbohydrate antigen 19-9 (CA19-9) were measured in 254 and 227 participants, respectively. Although the specificities of CEA and CA19-9 for CRC detection achieved approximately 90%, the sensitivities were pretty low (**Supplemental Table 5**).

## DISCUSSION

As one of the most common human cancers, the mortality of CRCs has been decreased with the emergence of new medical technologies and therapeutic agents. However, many patients still couldn’t be treated effectively or often suffered a low quality of life after the treatments. This partly attributed to their inability to be detected in precancerous lesions before progressing to cancerous or early-stage before spreading beyond the bowel wall. Several studies suggested that it should be suitable for CRC early detection because of the high correlation of mortality with disease stage ^23, 24^. Therefore, developing novel highly accurate, non-invasive and cost-saving tests is of critical importance for screening in large-scale population with average-risk of CRC ^25–27^.

Traditional diagnostic methods, such as colonoscopy and histopathological examination have been widely used for the detection of CRC. The remarkable sensitivity of colonoscopy made it the reference method for both cancerous and precancerous lesions screening, however obvious limitations such as the invasive procedures and low patient compliance are still existed ^28, 29^. Fecal occult blood test (gFOBT) and fecal immunochemical test (FIT) are two noninvasive screening methods based on occult blood in the stool, while both of them showed somewhat lower sensitivities and might lead to false-negative test results ^30^. Similarity was obtained for the serum markers of CEA and CA19-9 ^31^. The methylation-based, noninvasive stool DNA test has been documented many advantages with superior accuracy, as well as convenient procedures and high patient compliance. At present, many studies are focusing on developing novel methylated targets for CRC screening, despite that few of them are commercially available. Cologuard was the first commercially available MT-sDNA test approved by FDA and the sensitivity reached 92.3% ^7^. The other two commercially available sDNA tests were ‘EarlyTect-Colon Cancer’ (Genomictree, South Korean) and Colosafe (Creative Biosciences CO., Ltd., China), of which both were based on the aberrantly methylated SDC2, and their sensitivities were approximately around 90% ^9, 10^. Our findings showed that the dual-target sDNA test achieved an excellent sensitivity of 95.31% for CRC detection, outperforming not only the serum markers but also two commercially available sDNA tests. For advanced adenomas, it had a detection rate of 63.16%, which was higher than that of ‘Consafe’ and comparable to the ‘EarlyTect’. Besides, the DT-sDNA test showed superior sensitivity and specificity for the detection of both CRC and adenoma when compared to Epi proColon 2.0 ^13^.

It should be noted that combining two single targets, which means increasing the number of false positives, could decrease the specificity of the test. Our results showed that the specificity of this DT-sDNA test was lower than that of the two single-target sDNA tests, ‘EarlyTect’ and ‘Colosafe’. However, when using healthy normal controls and interfering diseases as negative controls separately, the specificities improved from 88.39% to 96.67% and 93.18%, respectively. This finding implies that the disadvantage could be eliminated or at less diminished by integrating clinical symptoms of participants during the testing.

Meanwhile, we did not observe significant variations of this DT-sDNA test in detecting CRCs with different clinical features, indicating its ability is not affected by patient gender and age, as well as tumor locations and differentiation. Moreover, the capabilities of SDC2 and TFPI2 alone in detecting left-sided and right-sided CRCs showed significant preference. Given the huge distinctive features of left- and right-sided colons ^22, 32^, it would be reasonable for the preference of the methylated SDC2 and TFPI2 in detecting different locations’ CRCs, which could elucidate, from one hand the improved sensitivity of combined two targets than that of any single target alone.

In summary, the performance of this novel dual-target stool DNA test for the diagnosis of CRCs was evaluated using large-scale data collected from a multicenter clinical trial. Outcomes of this study showed high sensitivities of the test in detecting CRCs and remarkable specificities were also observed in screening interfering diseases and healthy normal controls. However, some shortcomings of the test still exist. The sensitivity for adenomas detection is not yet pretty well, especially for the detection of non-advanced adenomas. In addition, the performance in screening asymptomatic persons at average risk of CRC needs to be assessed in a larger population. For screening purposes, the target for both sensitivity and specificity of a test would be not the same as diagnostic purposes ^33^. Nevertheless, the robustness of this non-invasive dual-target test in clinical practice indicated that it could be used as a screening option for individuals with average risk of CRC.

## Data Availability

All necessary data used in this study are presented in the manuscript and supplementary materials. The raw data are available from the corresponding author on reasonable request.

## Authors’ Contributions

Study design: YL Dou, J Lin, ZX Wang

Study supervision: YL Dou

Data collection (patient information, stool sample preparation, etc.): ZX Wang, S Jian, GN Zhang,

LJ Kong, F Zhang, Y Guo

Methodology development: S Jian,LJ Kong, F Zhang, Y Guo

Data analysis and interpretation (e.g., statistical analysis, computational analysis): GN Zhang, LJ Kong

Manuscript writing, review, and revision: YL Dou, J Lin, ZX Wang

Other (i.e. stool DNA extraction, methylation-specific PCR): GN Zhang, LJ Kong

## Acknowledgments

We thank Yang Zhang, Jiadan Xu, Yuanyuan Hu, Fubing Wang, Guangming Ye and Jun Xiao, for their assistance in preparing stool samples and collecting participants’ information used in the study. We declare that submitted manuscript does not contain previously published material, and are not under consideration for publication elsewhere. Each author has made an important scientific contribution to the study and is thoroughly familiar with the primary data. All authors listed have read the complete manuscript and have approved submission of the paper. The manuscript is truthful original work without fabrication, fraud or plagiarism.

