## Supplementary tables 1-5 for "Evaluating the clinical performance of a novel dual-target stool DNA test for colorectal cancer detection"

**Supplemental Table 1** Summary of the participants received colonoscopy or histopathological examinations.

| Group | Diseases | Colonoscopy | Histological | Total |
|---|---|---|---|---|
| Normal | | 300 | 0 | 300 |
| Interfering diseases | Anal cancer | 0 | 1 | 1 |
| | Appendix cancer | 1 | 1 | 2 |
| | Colon polypus | 1 | 72 | 73 |
| | Coloproctitis | 31 | 139 | 170 |
| | Diverticulum | 15 | 1 | 16 |
| | Esophagitis | 0 | 1 | 1 |
| | Gastric polyps | 11 | 1 | 12 |
| | Gastritis | 13 | 2 | 15 |
| | Hemorrhoids | 65 | 5 | 70 |
| | Liver cancer | 0 | 2 | 2 |
| | Stomach cancer | 0 | 6 | 6 |
| | Other | 18 | 10 | 28 |
| Adenoma | Non-advanced adenoma | 0 | 110 | 110 |
| | Advanced adenoma | 0 | 38 | 38 |
| CRC | | 0 | 320 | 320 |
| Total | | 455 | 709 | 1164 |

**Note:** if the participants received both colonoscopy and histopathological examinations, we only used the histopathological examinations to determine their disease status.

**Supplementary Table 2** Performance of methylated *SDC2* and *TFPI2* in detection of CRCs and other groups.

| Traget | Sensitivity (%) | Specificity (%) | AUC(95%CI) | Compared group |
|---|---|---|---|---|
| SDC2 | 71.74 | 96.2 | 0.84(0.81~0.86) | All non-CRC vs CRC |
| TFPI2 | 83.85 | 88.95 | 0.86(0.84~0.89) | All non-CRC vs CRC |
| Dual-target | 95.34 | 88.36 | 0.92(0.90~0.93) | All non-CRC vs CRC |
| SDC2 | 71.74 | 100 | 0.86(0.84~0.88) | Normal vs CRC |
| TFPI2 | 83.85 | 96.67 | 0.90(0.88~0.93) | Normal vs CRC |
| Dual-target | 95.34 | 96.67 | 0.96(0.94~0.97) | Normal vs CRC |
| SDC2 | 18.24 | 100 | 0.59(0.56~0.63) | Normal vs Adenoma |
| TFPI2 | 39.19 | 96.67 | 0.68(0.64~0.71) | Normal vs Adenoma |
| Dual-target | 41.22 | 96.67 | 0.69(0.66~0.73) | Normal vs Adenoma |
| SDC2 | 36.84 | 100 | 0.68(0.61~0.77) | Normal vs Advanced adenoma |
| TFPI2 | 57.89 | 96.67 | 0.77(0.70~0.84) | Normal vs Advanced adenoma |
| Dual-target | 63.16 | 96.67 | 0.80(0.73~0.87) | Normal vs Advanced adenoma |
| SDC2 | 11.82 | 100 | 0.56(0.53~0.59) | Normal vs Non-advanced adenoma |
| TFPI2 | 32.73 | 96.67 | 0.65(0.61~0.69) | Normal vs Non-advanced adenoma |
| Dual-target | 33.64 | 96.67 | 0.65(0.62~0.70) | Normal vs Non-advanced adenoma |
| SDC2 | 73.01 | 100 | 0.87(0.83~0.90) | Normal vs Stage I-II |
| TFPI2 | 81.6 | 96.67 | 0.89(0.87~0.92) | Normal vs Stage I-II |
| Dual-target | 96.93 | 96.67 | 0.97(0.95~0.98) | Normal vs Stage I-II |

| | | | | |
|---|---|---|---|---|
| SDC2 | 69.18 | 100 | 0.85(0.81~0.88) | Normal vs Stage III-IV |
| TFPI2 | 84.93 | 96.67 | 0.91(0.88~0.94) | Normal vs Stage III-IV |
| Dual-target | 93.15 | 96.67 | 0.95(0.92~0.97) | Normal vs Stage III-IV |

**Supplementary Table 3** Performance of the DT-sDNA test in discriminating interfering diseases from CRCs.

| Interfering diseases | Subject (%, n) | Positive detection rate (%, n) | Specificity (%, n) |
|---|---|---|---|
| Colon polypus | 18.53 (73) | 2.23% (9) | 87.67 (64) |
| Coloproctitis | 42.89 (169) | 2.48% (10) | 94.08 (159) |
| Diverticulum | 4.06 (16) | 0.25% (1) | 93.75 (15) |
| Esophagitis | 0.25 (1) | 0 (0) | 100 (1) |
| Gastric polyps | 3.05 (12) | 0 (0) | 100 (12) |
| Gastritis | 3.81 (15) | 0 (0) | 100 (15) |
| Hemorrhoids | 17.77 (70) | 0.99% (4) | 94.29 (66) |
| Liver cancer | 0.51 (2) | 0 (0) | 100 (2) |
| Stomach cancer | 1.27 (5) | 0.25% (1) | 80 (4) |
| Appendix cancer | 0.51 (2) | 0 (0) | 100 (2) |
| Anal cancer | 0.25 (1) | 0 (0) | 100 (1) |
| Other | 7.11 (28) | 0.5% (2) | 92.86 (26) |
| **Total** | **100 (394)** | **6.70% (27)** | **93.15 (367)** |

**Supplementary Table 4** Retested results of 34 resected patients by DT-sDNA test.

| | | Before resected | |
|---|---|---|---|
| | | Positive | Negative |
| After resected | Positive | 2 | 0 |
| | Negative | 28 | 4 |

**Supplementary Table 5** Performance of CEA and CA19-9 for CRC detection.

| | CEA | | | | CA19-9 | | | |
|---|---|---|---|---|---|---|---|---|
| | Negative | Positive | Sensitivity | Specificity | Negative | Positive | Sensitivity | Specificity |
| Healthy control | 88 | 11 | 0.29 | 0.89 | 74 | 7 | 0.12 | 0.91 |
| CRC | 112 | 45 | | | 131 | 17 | | |
